# Pleiotropic Genetic Effects between Multiple Sclerosis and Musculoskeletal Traits

**DOI:** 10.1101/2023.09.12.23295444

**Authors:** Sohyun Jeong, Ming-Ju Tsai, Changbing Shen, Yi-Hsiang Hsu

## Abstract

**Background:** Musculoskeletal disorders were commonly reported in patients with multiple sclerosis. However, the underlying etiology linking Multiple Sclerosis (MS) and musculoskeletal disorders is not well studied. With large-scale Genome-Wide Association Studies (GWAS) publicly available, we conducted genetic correlation analysis to identify shared pleiotropic genetic effects between MS and musculoskeletal traits. We also conducted Mendelian Randomization (MR) to estimate the causal relation between MS and increased risks of musculoskeletal disorders.

**Methods:** Linkage Disequilibrium Score Regression (LDSR) analysis was performed to estimate heritability and genetic correlation. Univariable, multivariable, and bidirectional MR analyses were conducted to estimate the causal relation. These analyses were done by utilizing the recent GWAS summary statistics of MS, fracture, frailty, falls, and several musculoskeletal risk factors, including bone mineral density, lean mass, grip strengths, and vitamin D.

**Results:** LDSR analysis showed a moderate genetic correlation of MS with falls (RG=0.10, *p=0.01*) but not with fracture and frailty. Genetic variants (rs13191659) in *LINC00240* gene which is associated with iron status biomarkers was found to be associated with both MS and falls. In MR analyses after excluding outlier SNPs with potential pleiotropic effects and correcting for multiple testing, MS presented no causal association with fracture and frailty but a minimal association with falls. Falls showed causally increased risks of fracture and frailty.

**Conclusion:** Our study suggests a potential genetic correlation with shared pleiotropic genetic effects between MS and falls. However, we didn’t find evidence to support the causal relation between MS and increased risks of falls, fracture, and frailty.

## Introduction

Multiple sclerosis (MS) is a demyelinating inflammatory autoimmune disorder presenting continuous and diffuse changes in the white and grey matter, breakdown of myelin, and damage to axons [1]. Despite recent progress characterized by the advent of new disease-modifying therapies, patients with MS still show an increased risk of musculoskeletal disorders such as fracture [2, 3], accidents and falls [4], and frailty [5] compared to controls. A meta-analysis comprising 9 cohort studies proposed significantly increased fracture risk in MS by 1.58 times compared to controls [6]. Falls are a debilitating consequence of MS and 56% of MS patients experience falls in any 3-month period [4]. Frailty is a marker of poor prognosis in patients suffering from systemic sclerosis [5] and a significantly higher percentage of MS patients were frail compared to controls (28% vs 8%) [5].

Multiple attributable risk factors of musculoskeletal disorders of MS have been proposed, including secondary causes delineating systemic bone loss by chronic inflammatory status [7], muscle weakness due to immobility [8], and ataxia from disruption of neuronal impulses [9], as well as bone and muscle loss due to MS treatment such as corticosteroids use [10], and psychotropic drugs (antidepressants, anxiolytics, and anti-epileptics) use [11, 12], which make MS patients vulnerable to falls. Beyond the secondary causes, intrinsic factors of MS etiology might be associated with the increased musculoskeletal disorders such that early stage comparatively young MS patients with minimal or no physical disability also presented low BMD [13].

However, most of the findings were typical observational studies not precluding potential biases from undefined residual confounding and reverse causation. Therefore, the elucidation of the effects of genetically unmodifiable predisposing factors will give further insights into preventative strategies for MS patients’ care.

In this context, LDSR is a reliable and efficient method of using GWAS summary-level data to estimate the genetic correlation between different phenotypes [14]. In addition, MR is a widely used method using measured variation in genes (genotypes, SNPs) of exposure to examine the causal effect of exposures on disease outcomes in observational studies [15, 16]. We implemented LD score regression as well as univariable, multivariable and bidirectional MR, and estimated genetic pleiotropic effect (correlation) and direct MS effects to 3 musculoskeletal outcomes (fracture, falls, and frailty) adjusting for the confounding effects from well-established bone and muscle disorder-related risk factors such as bone mineral density (BMD), lean body mass (whole body fat-free mass, appendicular lean mass), grip strengths and vitamin D.

We performed the following analyses: (1) LDSR to assess genetic heritability and genetic pleiotropy (correlations) between MS and musculoskeletal traits; (2) univariable MR to assess the causal effect of genetic determinants of MS on fracture, falls, frailty, and multiple risk factors; (3) multivariable MR to evaluate independent specific effects of MS to fracture, falls, frailty adjusting for other contributing risk factors; (4) bidirectional MR to assess the directionality among fracture, falls, frailty.

## Methods

### Linkage Disequilibrium Score Regression (LDSR)

We used the LD Hub interface which provides an automatic LD score regression analysis pipeline for users [17]. To standardize the input file, quality control is automatically performed on the uploaded file in LD Hub. In order to restrict the analysis to well-imputed SNPs, LD Hub filters the uploaded SNPs to HapMap3 SNPs [18] with 1000 Genomes EUR MAF above 5%, which tend to be well-imputed in most studies. In addition, to make the estimates of the genetic correlation to be reliable, the recent MS GWAS meta-analysis of the International Multiple Sclerosis Genetics Consortium (IMSGC) comprising 47,429 cases ad 68,374 controls [19] needs to meet the following criteria;1) Z score is at least > 1.5 (optimal > 4), 2) Mean Chi-square of the test statistics > 1.02, and 3) the intercept estimated from the SNP heritability analysis is between 0.9 and 1.1 [17]. We used UK Biobank GWAS datasets provided by LD hub pipeline estimating genetic heritability and correlation with MS.

### Mendelian Randomization (MR)

Supplementary Table 1 describes the detailed data source used in MR study. The largest publicly available GWAS summary statistics were used. Most GWAS summary statistics data were downloaded from the “MRC IEU OpenGWAS database” repository (https://gwas.mrcieu.ac.uk/) or each GWAS consortia. The overall study design for MR is displayed in Fig 1.

**Fig 1.**
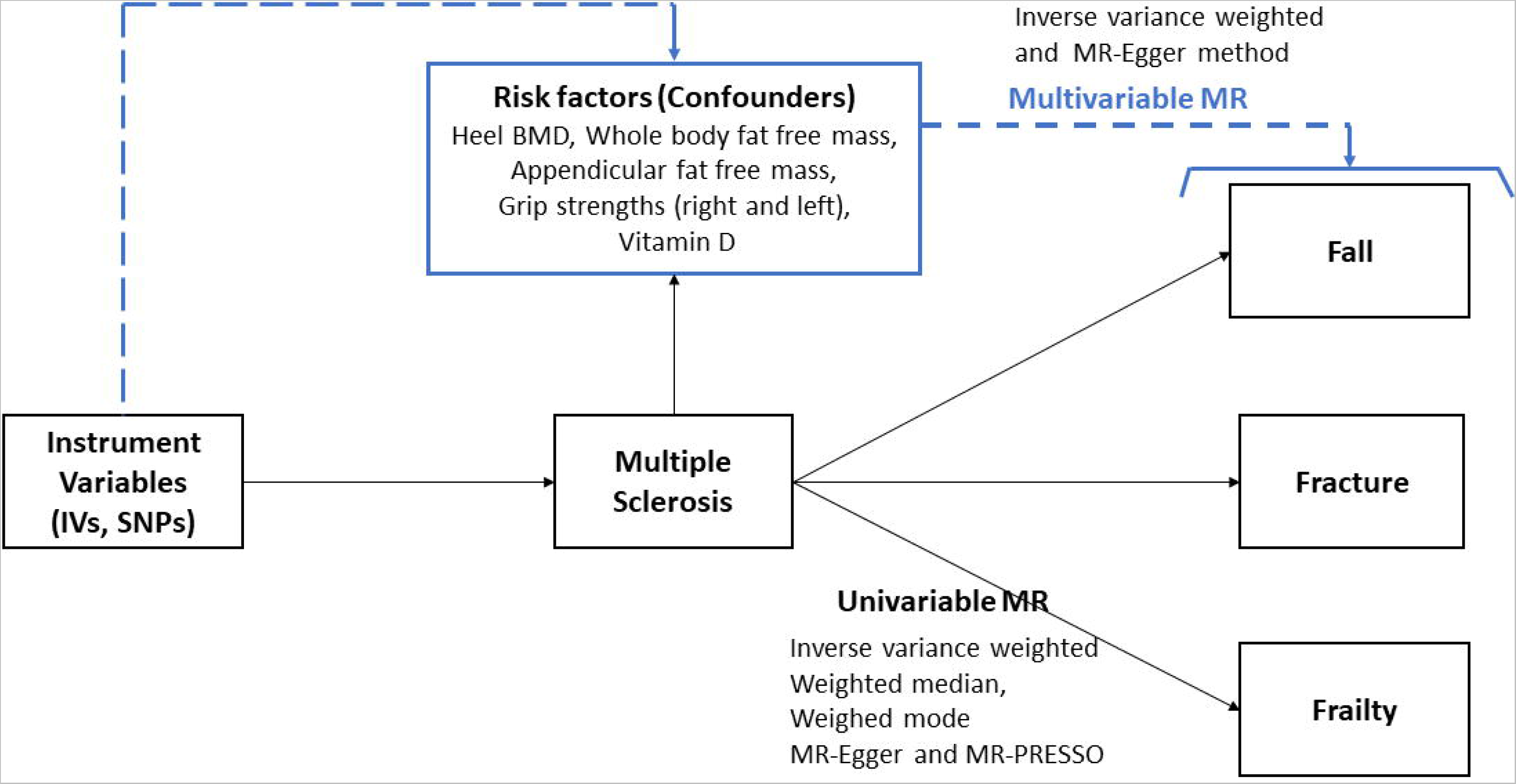
Conceptual model of univariable and multivariable Mendelian Randomization.

#### Exposure data

Instrumental variables (IVs) for MS are obtained from the recent GWAS meta-analysis of the IMSGC [19]. Among the 233 genome-wide statistically independent associations with MS susceptibility, 200 effects (SNPs) located in the autosomal non-MHC genome were selected as IVs (Supplementary Table 2). The detailed analysis and results description is provided in the previous IMSGC publication [19]. SNPs of genome-wide and suggestive effects jointly explain ~48% of the estimated heritability for MS [19].

#### Outcome data

Fracture and falls GWAS data used in the analysis were selected from the UK Biobank data, fractured/broken in last 5 years (ID: ukb-b-13346, n=44,502 cases and 415,887 controls) [20] and falls last year (ID: ukb-b-2535, n=461,725). Frailty GWAS data was from Atkins et al, 2019 (n=175,226) [21].

#### Risk factors of musculoskeletal outcomes

Known bone and muscle strength-related factors were used as covariates in MVMR. They are eBMD (heel)[22], appendicular lean mass, whole body fat-free mass, right handgrip strength, left grip strength [23] vitamin D [24], frailty [21], and falls [25].

### Statistical analysis

#### Univariable MR between MS and three musculoskeletal traits as well as risk factors

Univariable MR was applied to assess the causal relation between MS and 3 musculoskeletal outcomes (fracture, falls, and frailty) and 5 risk factors. Furthermore, MR effects of musculoskeletal risk factors on 3 musculoskeletal outcomes were assessed to examine the potential genetically determined risk paths of 3 musculoskeletal outcomes. Multi-instrument MR approaches were applied, which were to perform a single instrumental variable MR first; and then combined MR results from multiple instrumental variables via fixed-effect meta-analysis to estimate the overall causal relationships.

#### MVMR between MS and three musculoskeletal outcomes adjusting for risk factors

We also applied regression-based MVMR to account for many variants having pleiotropic effects that are associated with multiple risk factors in order to provide coefficients (effect sizes) representing the direct causal effects of MS to three musculoskeletal outcomes in turn with the other risk factors being fixed. MVMR is an extension of MR to deal with genetic variants that are associated with multiple risk factors. We selected risk factors that are known to be biologically related to musculoskeletal traits and where there is a potential network of causal effects (mediation) from one risk factor to another. The causal estimate is obtained by regression of the associations with the outcome on the associations with the risk factors, with the intercept set to zero and weights being the inverse-variances of the associations with the outcome [26]. Risk factors presenting significant univariable MR association with each musculoskeletal trait were included in MVMR.

#### Bidirectional MR among the three musculoskeletal outcomes

Bidirectional MR was performed among fracture, fall and frailty to examine their mutual effects and directionality considering their highly correlative nature. In bidirectional MR, instruments for both exposure and outcome are used to evaluate whether the “exposure” variable causes the “outcome” or whether the “outcome” variable causes the “exposure”[27].

#### Heterogeneity analysis

We used Cochran’s Q statistic to check the heterogeneity of IVs. Heterogeneity indicates a possible violation of the necessary IVs or modeling assumptions of which pleiotropy is a likely major cause. Cochran’s Q statistic derived from the inverse variance weighted (IVW) estimate should follow a χ^2^ distribution with degrees of freedom equal to the number of SNPs minus 1. Excessive heterogeneity is an indication that either the modeling assumptions have been violated, or that some of the genetic variants violate the IV assumptions—e.g. by exerting a direct effect on the outcome, not through the exposure [28]. This is termed ‘horizontal pleiotropy’[29].

When Q statistics showed significant heterogeneity, we applied MR-PRESSO [30] method to reassess the MR effects after filtering out outlier SNPs and compared the fixed MR results with IVW-based results.

#### Sensitivity analysis

The sensitivity analysis including IVW, weighted median, weighted mode, and MR-Egger tests were performed in univariable MR analysis. For MVMR, IVW and MR-Egger tests were evaluated. To exclude instrumental variables that may have horizontal pleiotropy and cause false positive findings of the causal inference, we also applied the MR-Egger regression test [31] to verify horizontal pleiotropy in each MR. Among the multiple methods, we set the results from IVW and MR-PRESSO outlier removal results as primary findings.

All the MR analyses were conducted in R (version 4.0.0) using TwoSampleMR [32], MendelianRandomization [33], MRPRESSO [34] packages, and the MR-Base platform [35]. The causal effect size (beta) with standard error and p-values were presented as appropriately. We applied a false discovery rate (FDR) to correct for multiple testing. The criteria used to select final instrument variables in each MR were GWAS significance: p<5e^-8^, LD clumping: r^2^>0.001 within a 10MB window, and proxy SNPs in 1000 Genomes EUR r^2^=0.8 [36].

## Results

### Genetic heritability and correlations by LDSR

Regression weight LD Scores for 1,293,150 SNPs of MS GWAS data were read. After merging with reference panel LD, a total of 1,109,876 SNPs remained for the final analysis. LDSR demonstrated that MS has statistically significant moderate genetic correlation with falls (rG: 0.105, *p=0.010*) but not with fracture (rG: −0.017, *p=0.711*) or frailty (rG: 0.151, *p=0.082*). Both handgrip strengths and both leg fat-free masses also showed a genetically significant correlation with MS. However, the whole-body fat free mass didn’t have a significant correlation with MS. The heritability of these traits estimated from GWAS summary statistics was low or considered not high; fracture (*h^2^*:0.019), falls (*h^2^*: 0.033), and frailty (*h^2^*: 0.015) (Table 1). We found genetic variant rs13191659 mapped to *LINC00240* (Long Intergenic Non-Protein Coding RNA 240) gene is commonly associated with MS and fall (MS GWAS p <5e-8 and fall GWAS<5e-5). *LINC00240* is known for total iron binding capacity [37], folic acid deficiency anemia and mean corpuscular volume [38].

**Table 1.** Genetic correlations between MS and musculoskeletal phenotypes and heritability by LD score regression.

| Trait | rG | SE_rG | Z_rG | P_rG | h2_obs | h2_obs_se | h2_int | h2_int_se |
| --- | --- | --- | --- | --- | --- | --- | --- | --- |
| Fractured/broken bones in last 5 years | -0.017 | 0.047 | -0.371 | 0.711 | 0.019 | 0.002 | 1.007 | 0.008 |
| Falls in the last year | 0.105 | 0.041 | 2.591 | **0.010** | 0.033 | 0.002 | 1.004 | 0.008 |
| Frailty | 0.151 | 0.087 | 1.741 | 0.082 | 0.062 | 0.028 | 1.056 | 0.009 |
| Heel bone mineral density (BMD) | -0.003 | 0.025 | -0.132 | 0.895 | 0.297 | 0.033 | 1.080 | 0.035 |
| Whole body fat-free mass | 0.035 | 0.025 | 1.395 | 0.163 | 0.291 | 0.012 | 1.114 | 0.029 |
| Hand grip strength (right) | -0.066 | 0.027 | -2.454 | **0.014** | 0.104 | 0.004 | 1.058 | 0.013 |
| Hand grip strength (left) | -0.067 | 0.028 | -2.427 | **0.015** | 0.104 | 0.004 | 1.055 | 0.012 |
| Leg fat-free mass (right) | 0.060 | 0.025 | 2.359 | **0.018** | 0.266 | 0.011 | 1.099 | 0.026 |
| Leg fat-free mass (left) | 0.062 | 0.025 | 2.435 | **0.015** | 0.268 | 0.011 | 1.092 | 0.025 |
| Arm fat-free mass (right) | 0.019 | 0.025 | 0.766 | 0.444 | 0.267 | 0.011 | 1.091 | 0.026 |
| Arm fat-free mass (left) | 0.030 | 0.025 | 1.191 | 0.234 | 0.267 | 0.011 | 1.097 | 0.025 |

rG: genetic correlation between two traits, SE\_rG: standard error of the genetic correlation, Z\_rG: Z score of the genetic correlation, P\_rG: p value of the genetic correlation. h2\_int: heritability intercept

### Mendelian Randomization

The univariable MR analysis showed a minimal causal effect of the total 109 MS SNPs to fracture, falls, and frailty in the IVW method. After excluding SNP outliers by MR-PRESSO and applying multiple testing corrections, without meaningful ORs, MS doesn’t seem to causally increase risks of falls (OR:1.004, 95% CI: 1.001-1.006, *q=0.018*); fracture (OR:1.002, 95% CI:1.000-1.003, *q=0.056*) and frailty score (beta:0.009, 95% CI: 0.001-0.017, *q=0.147)* and none of the univariable MR effects between MS and musculoskeletal risk factors were significant (Table 2 & Supplementary Table 3). Sensitivity analyses with different MR methods such as MR Egger, Weighted median, and Weighted mode methods didn’t replicate the significant results in all the MR analyses. All pleiotropy tests (MR Egger intercept test) were insignificant (Supplementary Table 3). Single SNP MR effects between MS and falls are presented in Supplementary Table 4 & 5. In MVMR analysis, except for adjustment of frailty, adjustment of each risk factor didn’t modify the overall MR results between MS and fall (Table 3). MVMR between MS and fracture as well as MS and frailty presented similar results that adjustment for each risk factor had no meaningful modification in overall MR results (Supplementary Table 6 & 7).

**Table 2.** Univariable MR effects between exposure (MS) and outcomes (musculoskeletal outcomes and risk factors)

| Exposure | Outcomes | MR effects between MS and outcomes |  |  |  |  |  |  |  |
| --- | --- | --- | --- | --- | --- | --- | --- | --- | --- |
|  |  | MR method | #SNP | beta | se | *p value* | Q value | OR | 95% CI |
| Multiple Sclerosis | Musculoskeletal traits |  |  |  |  |  |  |  |  |
|  | Fracture | IVW | 109 | 0.002 | 0.001 | 0.001 |  | 1.002 | 1.000-1.004 |
|  |  | MR-PRESSO* |  | 0.002 | 0.001 | 0.007 | 0.056 | 1.002 | 1.001-1.003 |
|  | Fall | IVW | 109 | 0.004 | 0.001 | 0.005 |  | 1.004 | 1.002-1.006 |
|  |  | MR-PRESSO |  | 0.004 | 0.001 | 0.002 | 0.018 | 1.004 | 1.001-1.006 |
|  | Frailty (continuous) | IVW | 152 | 0.012 | 0.004 | 0.007 |  | - | 0.004-0.020 |
|  |  | MR-PRESSO |  | 0.009 | 0.004 | 0.021 | 0.147 | - | 0.001-0.017 |
|  | Musculoskeletal risk factors |  |  |  |  |  |  |  |  |
|  | Heel BMD | IVW | 110 | -0.002 | 0.005 | 0.722 | 0.722 | - | - |
|  | Whole body fat free mass | IVW | 109 | 0.004 | 0.004 | 0.258 | 0.722 | - | - |
|  | Appendicular fat free mass | IVW | 110 | 0.006 | 0.007 | 0.402 | 0.722 | - | - |
|  | Grip strength(right) | IVW | 109 | -0.004 | 0.003 | 0.183 | 0.722 | - | - |
|  | Grip strength(left) | IVW | 109 | -0.002 | 0.003 | 0.389 | 0.722 | - | - |
|  | Vitamin D | IVW | 105 | -0.003 | 0.003 | 0.427 | 0.722 | - | - |

Abbreviation: IVW, inverse variance weighted; MR-PRESSO, Mendelian Randomization Pleiotropy RESidual Sum and Outlier
No horizontal pleiotropy was found in all analysis tested by Egger regression intercept test.
Heterogeneity test (Q test) was significant in all analysis except MS->fracture MR analysis.
To overcome the heterogeneity of SNPs effects, MR-PRESSO outlier tests were applied and outlier corrected results were presented along with results from IVW method.
\*No outlier detected

**Table 3.** Multivariable MR between MS and fall adjusting for musculoskeletal risk factor.

|  | MR effects between MS and fall |  |  |  |  |
| --- | --- | --- | --- | --- | --- |
|  | beta | se | pval | OR | 95% CI |
| **Unadjusted model** | 0.004 | 0.001 | 0.005 | 1.004 | 1.002-1.006 |
| Adjusted by frailty | 0.002 | 0.001 | 0.058 | 1.002 | 1.000-1.004 |
| Adjusted by<br>appendicular fat free mass | 0.004 | 0.001 | 0.004 | 1.004 | 1.002-1.006 |
| Adjusted by<br>whole body fat free mass | 0.004 | 0.001 | 0.004 | 1.004 | 1.002-1.006 |
| Adjusted by heel BMD | 0.004 | 0.001 | 0.004 | 1.004 | 1.002-1.006 |
| Adjusted by right grip strength | 0.003 | 0.001 | 0.011 | 1.003 | 1.000-1.005 |
| Adjusted by left grip strength | 0.003 | 0.001 | 0.008 | 1.003 | 1.000-1.005 |
| ed by all except frailty | 0.003 | 0.001 | 0.022 | 1.003 | 1.000-1.005 |
| Adjusted by all | 0.002 | 0.001 | 0.184 | 1.002 | 1.000-1.004 |

To account for the directional effects among the 3 musculoskeletal outcomes; fracture, fall and frailty, we conducted bidirectional MR. As expected, Fall causally increased fracture risk (OR:1.215, 95% CI: 1.098-1.345, *p=1.63E-04*) and frailty risk (OR: 4.870, 95% CI: 2.412-9.832, p*=1.01E-05*).

## Discussion

In this study, we found genetic correlation with shared pleotropic effects between MS and fall proven by LDSR and MR with minimal effect size. However, we don’t find meaningful causal relation to support epidemiological observation with increased musculoskeletal traits in MS patients. The MS doesn’t causally increase risks of musculoskeletal traits. Based on the univariable MR between MS and musculoskeletal risk factors, none of the well-known muscle and bone-related risk factors (whole-body and appendicular fat-free masses, both grip strengths, heel BMD and vitamin D) were causally affected by MS. In bidirectional MR among the three musculoskeletal outcomes, fall causally increased fracture and frailty risk.

Taking into all these findings together, we didn’t provide evidence to support that MS causally increased risks of neither musculoskeletal traits nor other musculoskeletal risk factors. The widely observed musculoskeletal disorders in MS patients in clinical studies might be more likely due to secondary factors that are associated with MS disease progression and treatment.

When we further explored SNPs commonly found in MS and fall with the criteria of GWAS/suggestive GWA significant variants, genetic variants mapped to a gene associated with iron biomarkers was found. Therefore, we could hypothesize MS and falls share pleiotropic genetic effects in iron biomarker pathways. It is well known that there is a correlation between anemia and falls, and specifically patients with lower hemoglobin levels are more likely to fall [39]. MRI and histological studies also suggest that iron levels are dysregulated in MS: iron accumulates in grey matter and is depleted in normal-appearing white matter [40].

Most of the previous observational studies predominantly reported high fracture risk in MS in comparison to age-matched controls. Dyn one study, Bazelier et al, 2011 [41] reported 3 fold increased hip fracture risk in patients with MS during 5 years of follow-up. However, their baseline fractures were higher in MS than age, sex-matched controls, furthermore the baseline falls history was higher in MS. Therefore, increased fractures observed in MS patients in this study might have been via increased fall risk in this population.

Falls are common in people with MS, with a large international data set demonstrating that 56% MS individuals fall at least once within 3 months period with 37% categorized as frequent fallers [4]. Notably, MS patients fall more frequently and experience more injurious falls, and are more likely to attribute their falls to tripping and distraction [42].

Medication use is a well-established risk factor for falls, either directly (affecting balance, attention, or muscle tone) or indirectly (as proxies of underlying conditions influencing the risk of falling; joint pain, arthrosis, and cardiovascular diseases among many others). A recent falls GWAS study examined the genetic correlation of falls with medication use and found medications such as opioids, anti-inflammatory and anti-rheumatic drugs, anilides, and drugs for peptic ulcer and gastro-esophageal reflux disease were in positive genetic correlations with falls [43]. Medication use associated with MS treatments and complications should be one of the strong risk factors for falls in this population. However, we are not able to evaluate such hypothesis based on our current study design.

Based on our study, genetic factors of MS might predispose MS patients to increased risk of falls by shared genetic pleiotropy but with minimal effects. Therefore, taking into account the secondary environmental or treatment-related factors such as careful medication use and incorporating stringent fall prevention strategy in patient care will mitigate the undesirable muscle and bone effects in MS patients.

We have a few novelties and strengths to mention. Given the highly increased availability of public GWAS data and identification of thousands of trait-associated loci in each trait, we could utilize LDSR and a multifaceted MR approach together and could elucidate the stepwise causal relations, and draw new perspectives on musculoskeletal traits in MS patients. We also illuminated genetically predisposed iron biomarker alteration might have a role in fall risk in MS patients. However, we have some limitations. Due to the low heritability of musculoskeletal traits and risk factors (Table 1), incorporating MR analysis might induce limited implications as well as GWAS data from these phenotypes might not highly reliable to elucidate any causal inference.

In conclusion, genetic association between MS and falls still stands, suggesting there are shared pleiotropic genetic factors between these two phenotypes. However, we didn’t establish a causal relationship between MS and multiple musculoskeletal traits which were reported by observational studies.

## Author contribution

Sohyun Jeong: Conceptualization, Data acquisition, Analysis, Draft and revision of manuscript

Ming-Ju Tsai: Data acquisition, Revision of manuscript

Changbing-Shen: Revision of manuscript

Yi-Hsiang Hsu: Conceptualization, Revision of manuscript

## Conflicts of Interest Statement

Sohyun Jeong, Ming-Ju Tsai, Changbing Shen, and Yi-Hsiang Hsu declare that they have no conflict of interest.

## Data Availability

All data produced in the present work are contained in the manuscript

https://gwas.mrcieu.ac.uk/

## Acknowledgement

The author(s) received no financial support for the research, authorship, and/or publication of this article. R01 grant number in the acknowledge section “R01AR72199”.

## Notes

### Competing Interest Statement

The authors have declared no competing interest.

### Funding Statement

This study was funded by NIH grant (R01AR72199).

### Author Declarations

This study only used publicly available datasets from consortium or public GWAS repository. Entierly from MRC IEU OpenGWAS database repository (https://gwas.mrcieu.ac.uk/)

